# Collaborative Federated Learning behind Hospitals’ Firewalls for Predicting Histological Response to Neoadjuvant Chemotherapy in Triple-Negative Breast Cancer

**DOI:** 10.1101/2021.10.27.21264834

**Authors:** Jean Ogier du Terrail, Armand Léopold, Clément Joly, Constance Beguier, Mathieu Andreux, Charles Maussion, Benoît Schmauch, Eric W. Tramel, Etienne Bendjebbar, Mikhail Zaslavskiy, Gilles Wainrib, Maud Milder, Julie Gervasoni, Julien Guérin, Thierry Durand, Alain Livartowski, Kelvin Moutet, Clément Gautier, Inal Djafar, Anne-Laure Moisson, Camille Marini, Mathieu Galtier, Guillaume Bataillon, Pierre-Etienne Heudel

## Abstract

Triple-Negative Breast Cancer (TNBC) is a rare cancer, characterized by high metastatic potential and poor prognosis, and has limited treatment options compared to other breast cancers. The current standard of care in non-metastatic settings is neoadjuvant chemotherapy (NACT), with the goal of breast-conserving surgery and for an *in vivo* assessment of chemosensitivity. However, the efficacy of this treatment varies significantly across patients, and this histological response heterogeneity is still poorly understood partly due to the paucity of available curated TNBC data. Motivated by this problem, we investigate the use of machine learning (ML) to predict at diagnosis the histological response to NACT for early TNBC patients. To overcome the known biases of related small scale studies while respecting data privacy, we conduct, for the first time, a TNBC study in a multi-centric fashion behind hospitals’ firewalls using collaborative Federated Learning (FL). Thereby allowing access to enough TNBC data to sustain a complete response heterogeneity investigation. We show evidence that local ML models relying on Whole-Slide Images (WSIs) at diagnosis are able to predict the histological response to NACT as accurately as current clinical approaches, which rely on time-consuming expert annotations. We demonstrate that collaborative training further improves performance over single-center training outperforming clinical methods. Our ML model is interpretable by design, and we show that it is sensitive to specific histological patterns. While we identify known predictive biomarkers among them, this proof of concept for real-world collaborative FL paves the way for future biomarker discovery using unprecedently large datasets.

## 2 Introduction

Among all breast cancers, Triple-Negative Breast Cancer (TNBC) is one of the most aggressive, with poor prognosis and high risk of neoplastic relapse after treatment even in non-metastatic cases [1]. TNBC is defined by the absence of expression (defined in this work as less than 10% expression following the recent clinical guidelines [2, 3, 4]) of both estrogen receptor (ER), progesterone receptor (PR) and Human Epidermal growth factor Receptor 2 (HER-2) [5, 6]. As a consequence, TNBC is not sensitive to new effective systemic therapies [7] such as endrocrine therapy or HER2-targeted treatments, which, combined with widespread screening [8], have increased the 5-year survival rate of non-metastatic breast cancers to 91% [9] (a 21% increase from 75% measured in 1975 [9]). Furthermore, TNBC comprises a multitude of molecular subtypes difficult to differentiate at diagnosis, and linked to different prognosis [10]: Basal-like and Immunomodulatory respond generally better than Luminal Androgene Receptor (LAR) and Mesenchymal-like cancers. Therefore, as displayed in Fig. 1 (a), most early TNBC patients get a neoadjuvant chemotherapy (NACT) based on anthracyclines and taxanes [11] followed by breast-conserving surgery.

**Fig. 1.**
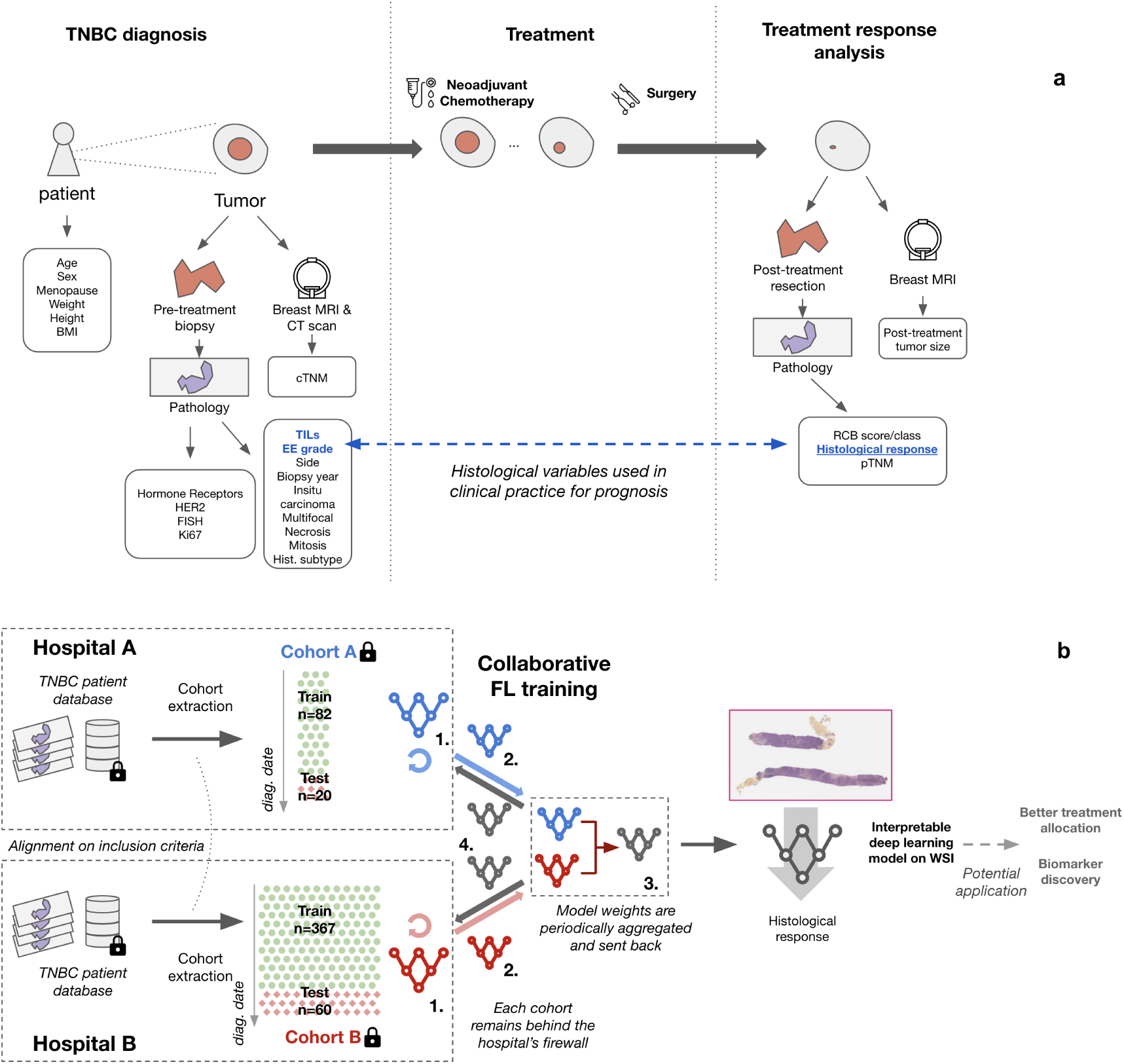
TNBC Treatment Pathway and Inter-Hospital Collaborative Federated Learning Study Overview. **(a)** Triple-Negative Breast Cancer (TNBC) treatment pathway. Left panel: TNBC is diagnosed following a primary tumor biopsy and subsequent histopathology analysis, breast MRI and CT-scan. These measures help derive demographic, clinical and histopathological variables describing the patient state at diagnosis. Middle panel: Treatment consists in neoadjuvant chemotherapy (NACT) followed by surgery. Right panel: Treatment response is analysed with another breast MRI and histological analysis, which help derive post-treatment TNM scores, the RCB and ultimately the histological response. Predicting histological response based on diagnosis variables is a difficult problem: standard clinical practice mostly relies on pre-treatment tumor Elston and Ellis (EE) histological grade and Tumor Infiltrating Lymphocytes (TILs) variables to guess the outcome. **(b)** Inter-Hospital Collaborative Federated Learning (FL) Study Overview. Two cohorts A and B were extracted from two different hospitals with similar inclusion criteria and format, with clinical variables described in (a) as well as pre-treatment biopsy whole slide imaging (WSI). A deep learning model was then trained in a Collaborative FL fashion, while keeping data behind each hospital’s firewall, so as to predict histological response based on WSIs. In Collaborative FL, models are trained locally (1), then sent to an aggregator node (2) where they are averaged (3) and the resulting consensus model is sent back to the workers (4), iteratively over multiple such rounds (1)-(4). The resulting model can predict NACT response from WSIs, and is interpretable by design. Potential future applications of this proof of concept include improved treatment allocation and biomarker discovery.

NACT response in early TNBC varies significantly across patients, but is not well understood. NACT response is often evaluated by the Residual Cancer Burden (RCB), a histological measure which has been shown to be directly correlated with disease free survival and overall survival [12]. In this work, we follow clinical practice and assess response to NACT by thresholding the resulting RCB class, giving rise to two classes of responders. Following clinical practice we count a positive Complete Response (pCR) if and only if RCB class is 0 after treatment, otherwise we consider the response to be incomplete, see details in Supp. Mat. B, Tab. 4. Being able to infer NACT response at diagnosis could help choose adequate treatments for patients earlier. For potential good responders, one could for instance de-escalate the chemotherapy by removing anthracyclines, thereby reducing the risk of cardiac toxicity. For potential non-responders, one could start other chemotherapy agents, such as carboplatin [13], or funnel patients towards clinical trials with more experimental treatments such as Poly ADP-Ribose Polymerase (PARP) inhibitors [14] or immunotherapies [15]. Understanding NACT response in early TNBC could therefore significantly impact treatment choice and ultimately improve survival.

The scientific community has recently started to draw connections between variables available at diagnosis and NACT histological response. While age at diagnosis seems to influence response [16, 17], there is also a growing consensus around the role of some histological variables, which are obtained through time-consuming expert annotations and sometimes Immunohistochemistry analysis (IHC). This is notably the case for the percentage of tumor-infiltrating T-lymphocytes (TILs) [18, 19, 20, 21] and the fraction of KI67–positive cells [22, 23, 24, 25, 16, 26, 17], and, less importantly, the Elston and Ellis (EE) histological grade [18, 16], and the presence of lymphovascular involvement or necrosis [27, 18, 28]. To date, none of the predictors mentioned has led to actionable classification methods at diagnosis for subtypes. Thus, in current clinical practice, oncologists and pathologists rely on practitioner-dependent and not formalized heuristics, mostly based on TILs and EE grade, in order to initiate NACT. Despite this progress, more breakthroughs are needed in order to achieve a complete understanding of NACT response.

Machine Learning (ML) on Whole-Slide Images (WSIs) is a promising approach to tackle open questions in oncology [29]. In other cancers, Deep Learning (DL) on WSIs has recently allowed the production of automatic extraction of clinical-grade known biomarkers [30, 31] and even to discover novel predictive biomarkers such as for Mesothelioma [32] or for Hepatocellular Carcinoma [33]. In the case of breast cancer, multiple works have investigated DL for breast cancer grading [34], prognosis [35] or even hormonal receptor status determination [36]. In the same line of work, it has been recently shown that TILs can be automatically detected in WSIs with high accuracy [37, 38], and that RCB can be computed on post-treatment WSIs with a DL model [39].

To the best of our knowledge, a single work has investigated the use of ML for NACT response prediction based on WSIs [40], and shown that WSIs contain useful signal for NACT response prediction. Yet, this study only reaches the same accuracy as clinicians, and relies on a relatively small dataset of 122 WSIs from a single center. The potential of ML to improve prognosis and discover new predictive biomarkers has yet to materialize in the case of NACT response in early TNBC.

Due to the relatively small prevalence of TNBC in the general breast cancer population (15% or 9, 000 new cases per year in France [41]), TNBC data is difficult to access in large quantities. The vast majority of published studies showcases very limited sample sizes, as historical data within each hospital is very limited, even in specialized cancer centers, see the comprehensive bibliographic search in Supp. Mat., Fig. 6 where the median patient count is 119 and 25%-75% quantiles are 82-207.

Collaborative Federated Learning [42, 43] is a promising novel paradigm allowing to perform multi-centric analyses without pooling data in a central repository. By preserving data control and ownership for their original institutions while enabling collaboration, this approach could revolutionize biomedical research, especially for relatively rare diseases like TNBC, by reaching a critical mass of data sufficient to train highquality ML models. However the complexity of its deployment in a real-world network of hospitals has hindered its progress, and so far federated learning studies have mostly been limited to simulations [44, 45, 46, 47, 48, 49, 50].

In this study, through an immense interdisciplinary effort across multiple institutions, we manage to perform the first real-world Federated Learning analysis on TNBC data, reaching for one of the first times a sample size of more than 500 TNBC patients between two hospitals (top 10% of studies in terms of patient counts according to Fig.6 in Supp. Mat.), which represents a milestone in TNBC research and more generally in showing the applicability of FL to biomedical research. Our main contributions are:

- Performing the first real-world collaborative Federated Learning between histopathology datasets behind hospitals’ firewalls, without ever sharing data itself, as displayed in Fig. 1 (b);
- Assembling one of the largest TNBC cohorts ever studied, with more than 500 TNBC patients from 2 institutions;
- Improving upon clinicians’ performance for predicting NACT response with DL models using only raw histopathology slides;
- Showing improved predictive power of Federated Learning models with respect to locally-trained models.

In addition to these four main contributions, we open the black-box of our best Federated DL model thanks to its interpretable architecture design, and discuss several new biomarkers hypotheses associated with NACT response in TNBC.

## 3 Results

### 3.1 Diagnosis clinical and histological variables and NACT response

We first examine the relationship between all available histological and clinical variables at diagnosis and NACT response independently in each center, relying both on univariate and multivariate analysis. The purpose of this investigation is to check the predictive power of known biomarkers, to provide a baseline score, and to gather insights on the specificities of each cohort.

#### Univariate Analysis

We first investigate individual variable relationship with NACT response in both centers. Fig. 2 (a) displays the associations between the available clinical variables and NACT response computed independently on each center, as measured through the Area Under the Curve of the Receiving Operator Characteristic (AUC). In Cohort A, we note a strong positive association with TILs: a large AUC is obtained using the variable itself to predict the NACT response (0.72 ± 0.12 AUC, *p*-value 5.98 · 10^−3^). No other variable in Cohort A demonstrates significant association with the histological response, including the EE histological grade despite its frequent use in clinical practice (0.51 ± 0.047 AUC, *p*-value *>* 0.05). In Cohort B, the fraction of KI67–positive cells has a significant relationship with response to NACT (0.59 ± 0.058 AUC, *p*-value 1.59·10^−2^), whereas TILs does not stand out from the univariate AUCs. These differences showcase the variability in the conclusions that can be drawn from TNBC studies with single-center cohorts.

**Fig. 2.**
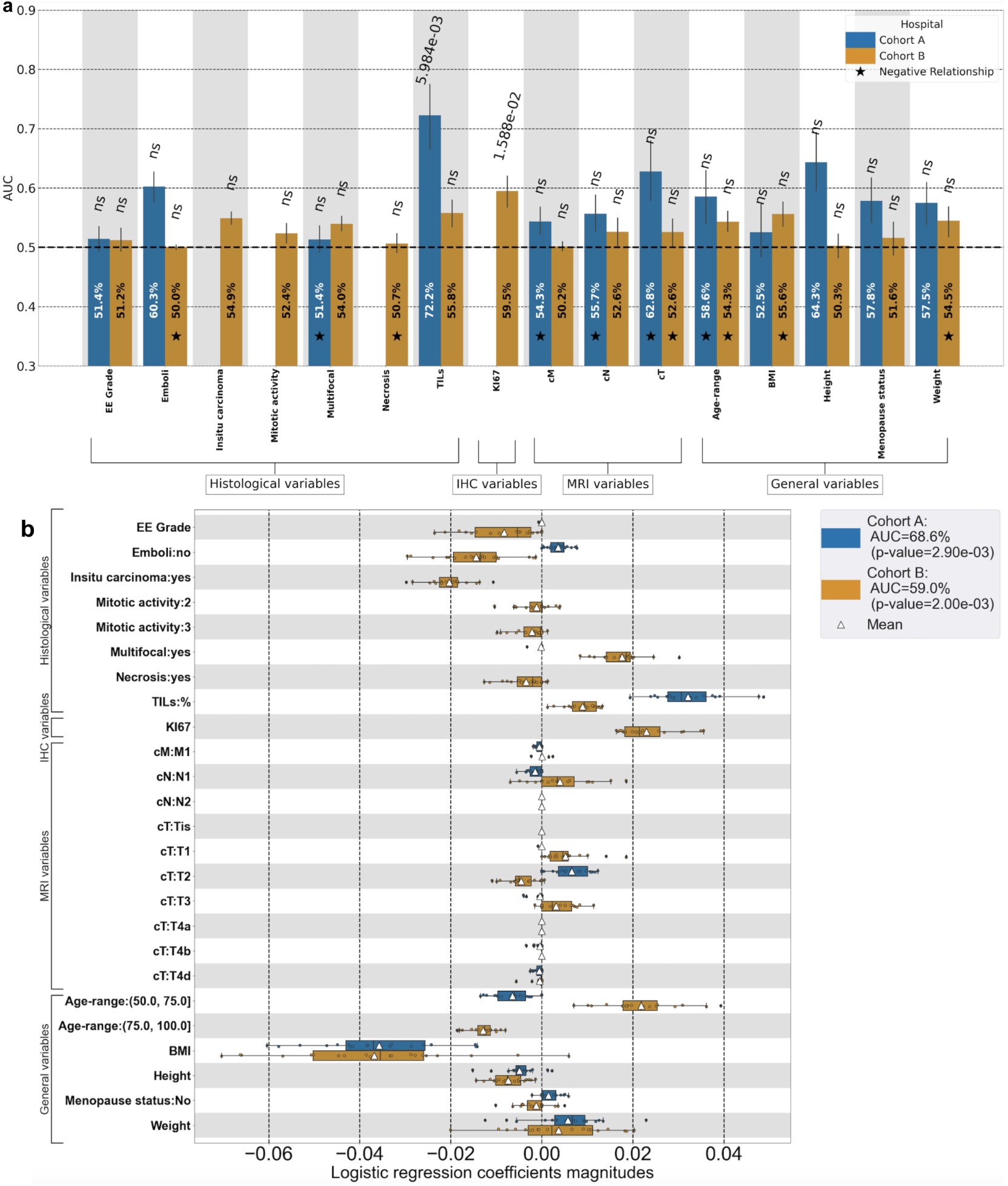
Univariate and Multivariate Association between diagnosis variables and NACT histological response. **(a)** Univariate association between raw values of variables collected at diagnosis and the NACT histological response for cohorts A and B, measured with the Area under the Curve of the Receiving Operator Characteristic (AUC). For each variable, AUCs are computed on each train set through five repeated 4-fold cross-validations: the bar’s height denotes the mean AUC and the error bar the 95% confidence interval over the resulting 20 values, while *p*-values are computed with respect to an unassociated random baseline. In Cohort A, only TILs is significantly associated with NACT response (*p*-value 5.99 · 10^−3^), while in Cohort B, only the fraction of KI67-positive cells is found significantly associated (*p*-value 1.59·10^−2^). **(b)** Amplitude of the coefficients of a logistic regression with ElasticNet penalty trained to predict NACT histological response from the collected variables on each individual cohort independently. Categorical variables are dummified. Dots represent individual values, boxplots represent their distribution (see Methods, Sec. 5), and the triangle denotes the mean. Despite the extensive collection of variables from different modalities, the NACT response prediction is volatile: very few variables can be clearly associated with the response and their effects vary with the cohort under study.

#### Multivariate Analysis

In order to refine the analysis and take into account interactions between variables, we independently conduct on each Cohort A multivariate logistic regression analysis with ElasticNet penalty [51] to predict NACT response, and check the average magnitude of the coefficients for each variable. We see in Fig. 2 (b) that univariate analysis results are confirmed: TILs and KI67 remain strong predictors with a mean magnitude of 0.03 and 0.022 on Cohort A and B respectively. Patients’ age category appears to have a stronger effect on Cohort B than what was observed in the univariate analysis, with a mean positive weight of 0.021 on the (50 *-* 75] age category on this cohort. We explain this result by the fact that older patients (*>* 75 years old) only appear in Cohort B, as detailed in Supp. Mat. C, and generally respond worse to NACT, leading to a difference in the weights attributed to the age variables between both cohorts. This experiment also allows, to some extent, to disambiguate for some variables the direction of association with the response, either positive or negative. For instance, Body Mass Index (BMI) seems to negatively influence NACT response in both cohorts, while for some variables like the age range, the effect varies between both cohorts.

Through this univariate and multivariate analysis, while we identify some of the clinical variables which are used in clinical practice such as TILs and KI67, we show some differences in predicting power of other variables between cohorts, which hint at differences between the cohorts themselves. We provide in the Supp. Mat. B an overview of each cohort in Tab. 3 as well as a complete description of the variables available in each Cohort and how they are measured in Tab. 4. Further, we provide in Supp. Mat. C a more complete discussion of the differences in the cohorts’ populations.

### 3.2 WSI-based models to predict NACT response

We now explore the impact of DL and Collaborative FL in predicting NACT response in early TNBC. We first compare the performance of clinical baseline models with DL trained on WSIs. We then investigate the impact of Collaborative FL on WSI-based DL models’ performance. The analysis relies on cross-validation and validation on held-out test data, as shown in Fig. 3 (a)-(b).

**Fig. 3.**
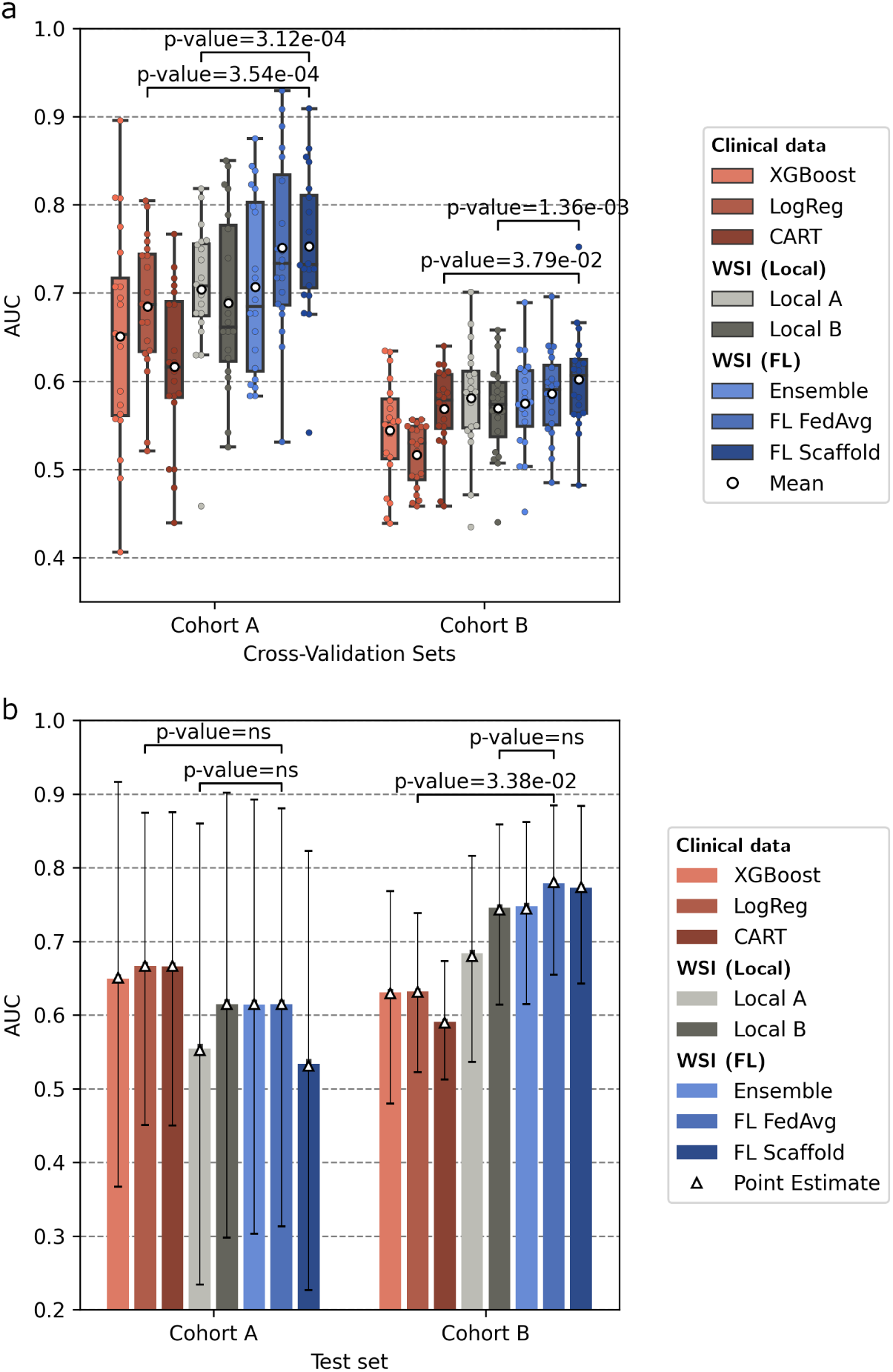
Performance comparison between local and collaborative FL training on clinical or WSI data to predict NACT treatment response in TNBC. **(a-b)** Performance comparison between the following trained models: XGBoost, Logistic Regression and CART trained locally on each cohort on tabular histological data, locally trained WSI-based model, an ensemble of previously trained local WSI-based models, and WSI model collaboratively trained with FederatedAveraging and Scaffold strategies. Models trained on tabular histological data on Cohort A (resp. Cohort B) are only tested on Cohort A (resp. Cohort B). Models trained on WSI data are separately tested on both cohorts. **(a)** Comparison of cross-validation performances between the different training strategies. DL models trained on WSIs with collaborative FL significantly outperform the best local DL WSI-based model as well as the best clinical baseline on both cohorts. Details on the boxplot parameters are available Sec. 5, Methods. **(b)** Comparison of performances between models trained on the entire training datasets and separately tested on the sanctuarized test set of each cohort. Overall the best Federated WSI-based model (FL FedAvg) leads to improved performances with respect to local trainings on both cohorts. More precisely, in Cohort B, DL models on WSIs trained with collaborative FL significantly outperform the clinical baseline.

#### Cross-validation results

We first compare the performance of single-center WSI-based DL models with clinical baseline models through cross-validation. Clinical baseline models are machine learning models trained independently on each center with two variables, EE histological grade and TILs, which are the clinical variables used in clinical practice. Since medical practitioners’ heuristics are not well defined and practitioner-dependent, we use these clinical baseline models to emulate them. Details on the models and methodology are provided in Sec. 5.3.2. In Fig. 3 (a), we note discrepancies ranging in 0.1 absolute AUC points between the local performances in both cohorts. We refer the reader to Sec. 3.1 and Supp. Mat. C to find a more complete discussion on the possible factors explaining the difficulty of the task on Cohort B. On Cohort A, locally trained WSI-based models reach a performance of 0.70 ± 0.079 AUC, which is on par with locally trained logistic regression of 0.68 ± 0.079 AUC (comparison *p*-value *>* 0.05). Similarly on Cohort B, locally trained WSI-based models reach a performance equivalent to that of the best clinical model (0.57 ± 0.053 AUC vs 0.57 ± 0.048 AUC, *p*-value*>* 0.05). Overall, cross-validation results show that local WSI-based DL models are able to reach a similar performance to the best models trained on expert-annotated data (TILs and EE histological grade), whose goal is to emulate the practitioner’s decision process.

We further investigate the ability of the WSI-based DL models to transfer between cohorts, as shown in Fig 3 (a). Local WSI-based DL models trained on Cohort B, where the task is harder, transfer well to Cohort A, reaching 0.69 ± 0.099 AUC (comparison *p*-value *>*0.05 with local training of 0.70 ± 0.079 AUC). Similarly, models trained on Cohort A transfer well to Cohort B, reaching a score close to the local training on Cohort B (0.58± 0.062 AUC vs 0.57± 0.053 AUC with comparison *p*-value *>* 0.05). These results indicate that locally-trained WSI-based DL models transfer well between both cohorts.

We now investigate the effect of collaborative training across cohorts on performance of WSI-based DL models. We use three collaboration strategies: Ensemble [52], which is a simple collaboration baseline with a single communication step, and two more complex FL strategies: FedAvg [42], and Scaffold [53]. We refer to Sec. 5.3.2 for more details. Results are depicted in Fig 3 (a). The Scaffold strategy is the most promising collaborative strategies, with 0.60 ± 0.056 AUC on Cohort B, which is significantly better than the locally-trained WSI-based DL model (AUC 0.57 ± 0.053, comparison *p*-value 1.36 · 10^−3^), and with 0.75 ± 0.083 AUC on Cohort A, which is significantly better than the corresponding local DL model (AUC 0.70± 0.079, *p*-value 3.12. 10^−4^). We also see that among distributed methods, Federated Learning methods perform generally better than ensembling local models. Further, the best federated results on both cohorts significantly outperform clinical baselines using TILs and EE grade (*p*-values 3.54 · 10^−4^ and 3.79 · 10^−2^ in Cohorts A and B, respectively) and is 0.03 points better on average than the strong TILs only univariate baseline on Cohort A (comparison p-value *>* 0.05). On Cohort B, the federated models are even on par with using expensive IHC annotation of KI67 Fig. 2 (a), KI67 obtains 0.60 ± 0.045 AUC, non-significant *p*-value. Cross-validation results therefore indicate that, on both centers, collaborative FL training improves performance of WSI-based DL models with respect to single-center training as well as clinical baseline.

#### Held-out results

We now investigate the generalization of the aforementioned models, training on the full cohorts and testing *once* on sanctuarized test sets from each cohort that were previously made inaccessible by their respective institutions of origin. On both cohorts, the best federated model slightly outperforms the locally-trained WSI-based DL models, although not significantly due to the small sample size. Indeed, on Cohort A, the local WSI-based DL model gives 0.55± 0.16 AUC, whereas the best FL model gives 0.62 ± 14.6 AUC (*p*-value *>*0.05); similarly, on Cohort B, the local model gives 0.74 ± 0.063 AUC and the best FL model 0.78± 0.059 AUC (*p*-value*>*0.05). When compared to clinical models on Cohort A, where TILs have been found to be very predictive, the best FL WSI-based model gives comparable performance to the best clinical model: the bootstrap test is not significant. On Cohort B, the best FL WSI-based model is significantly better than the best clinical model 0.78± 0.059 AUC vs 0.63± 0.055 AUC (*p*-value 3.38 · 10^−2^). Somewhat surprisingly, the performance gap is reversed and the model performances on the larger Cohort B is better than on Cohort A. We hypothesize that one reason is simply the variance of a point estimate on small sized test sets, as Cohort A’s test contains only 20 slides.

To synthesize, collaborative FL training provides a performance which is not significantly different from clinical baseline in Cohort A, and significantly improves upon clinical baseline in Cohort B. Confusion matrices of the models on the test sets can be found in the Supp. Mat., Fig. 7.

### 3.3 Interpretability of WSI-based model

One of the characteristics of our DL model is its ability to individually score each tile according to its response prediction. By design, the score of a patient is the average of the score of each tile. This allows us to extract the extremal tiles that contributed the most in a specific direction to analyse common patterns and try to understand the underlying biological patterns used by the model.

We extract the best federated model from Sec. 3.2, and use it to visualize, for each test slide, the individual tiles scores as obtained from the model prediction. The exact process to go from WSI score to tile score is described in Sec. 5.5. In addition to inspecting the scores distribution of the tiles of two representative patients (good and bad responder), we also plot in Fig. 4 (a) the distribution of the tiles score on the entire distributed dataset and extract the top and minimum scoring tiles across the slides from both centers. We show that the scores distribution on all tiles is bimodal, and that tiles associated with high and low scores are presenting clear histological patterns.

**Fig. 4.**
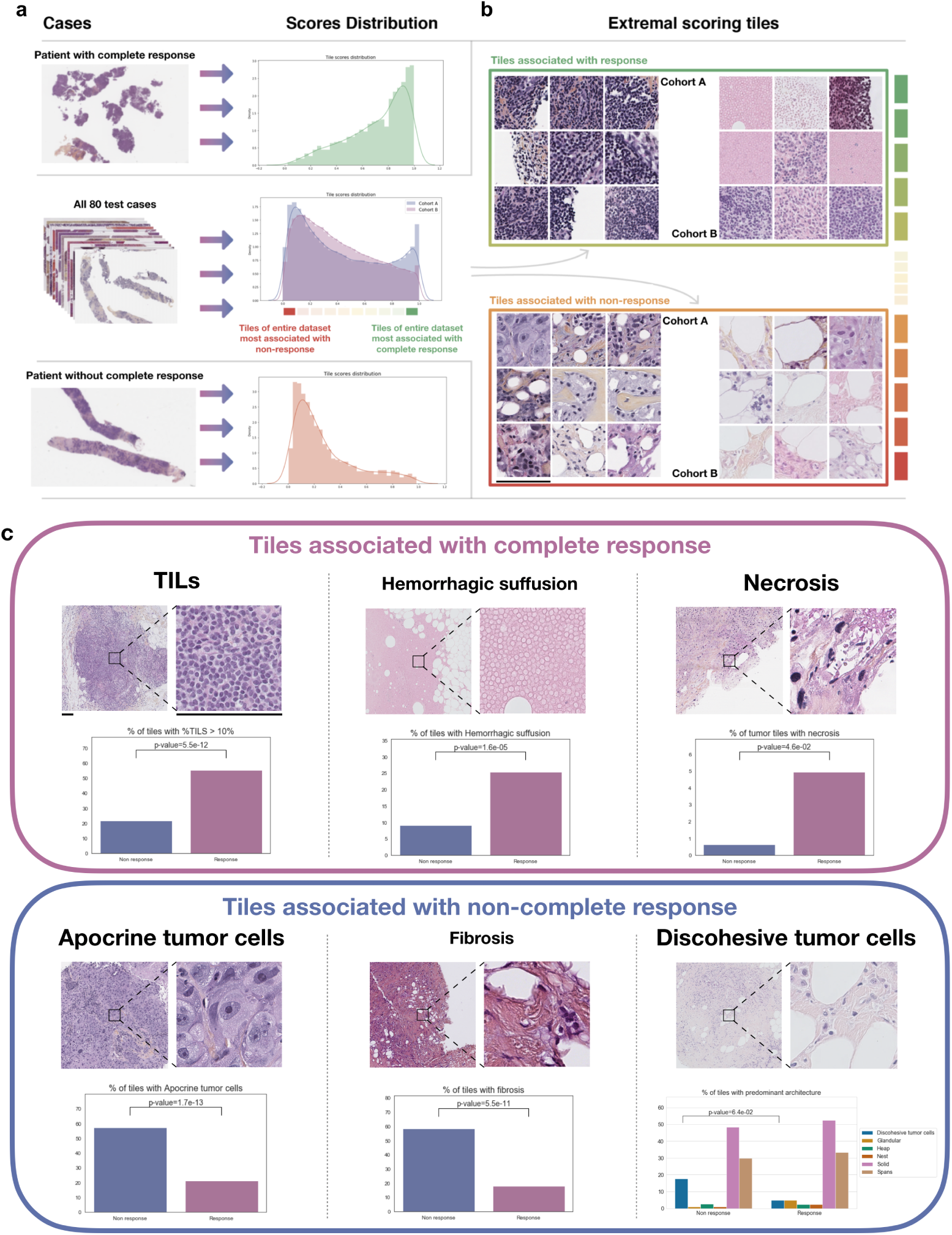
Interpretation of the WSI-based collaborative FL model. **(a)** The distribution of the tile scores obtained from raw WSIs using our FL model for a patient with complete response is shown on the top and without complete response on the bottom. In the middle, we show the aggregated score distributions for all tiles of all patients in the test sets (cohorts A and B). **(b)** Extremal tiles associated with response and no response for all TNBC patients were extracted to be reviewed by two pathologists. Scale bars, 112*µ*m **(c)** Examples of tiles classified as response or non-response by our model. Two expert pathologists blinded to risk scores analyzed the 200 most predictive tiles. The features predictive of response included TILs presence, hemorrhagic suffusion and necrosis. The features predictive of absence of response included presence of apocrine or discohesive tumor cells and fibrosis. Scale bars, 112 *µ*m.

Thanks to an extensive expert annotation of pre-defined criteria available in Tab. 5 in Supp. Mat., we are able to highlight specific pattern associated with response or non-response in the entire test sets. On Fig. 4 (b) we show that such patterns indeed appear clearly in the annotated tiles: presence of TILs (*p*-value 5.5 · 10^−12^), hemorrhagic suffusion (*p*-value 1.6 · 10^−5^) along with necrosis are associated with overall high scores and probability to respond, while presence of apocrine tumor cells with abundant eosinophilic granular cytoplasm and prominent nucleoli (*p*-value 1.7 · 10^−13^), fibrosis (*p*-value 5.5 · 10^−11^) and discohesive tumor cells are associated with low scores and high risk of poor NACT response. Most of those patterns are already well-known in the community (TILs, apocrine or discohesive tumor cells, hemorrhagic suffusion for example) and therefore confirm the biological relevance of our algorithm. Some other patterns, like the presence of fibrosis, require a deeper analysis to understand the link made by the algorithm between this pattern and a low probability of response.

## 4 Discussion

In this study, we show promising results for the applicability of WSI-based DL models in non-metastatic TNBC diagnostics, thanks to the availability of a secure federated network between comprehensive cancer centers. We now discuss the significance of these findings.

Our work presents one of the first use cases of *real-world* Federated Learning and Federated Analysis tackling open problems with rare diseases. Indeed, the vast majority of published Federated Learning studies [44, 45, 46, 47, 48, 49, 50] is analyzing data with synthetic splitting: authors first pool data and then *a posteriori* distribute it into simulated centers, either identified through metadata or with a synthetic split. This makes it considerably simpler to perform the analysis, and to artificially increase the number of centers.

To the best of our knowledge, our study is pioneer insofar as data is accessed behind hospitals’ firewalls: it is impossible for us to pool data due to contractual and security reasons, and data splits directly stem from these constraints. In spite of such stringent security requirements, we manage to conduct a distributed data analysis, while sharing only non patient-specific aggregated information, leading to novel insights.

In addition to pioneering this new setting, we purposefully chose a challenging task where there was only little evidence [40], prior to this work, that DL methods would be effective at all, instead of studying tasks such as direct TILs prediction from slides [37, 38], where the success of DL methods was to be expected given sufficient data. This work proves that Federated Learning can provide new insights on open problems by leveraging a joint analysis of data stored in each silo to better probe the underlying complexity of the scientific question, thereby ultimately maximising impact on patients’ lives.

More precisely, results on clinical data Fig. 2 both confirm clinical practice to measure TILs, and KI67 when available. EE histological grade is not significantly associated to NACT response in our results, but this is probably due to the cohorts being mostly composed of grade 3 patients, as shown in Supp. Mat. C. We show WSI-based models match or outperform the predictivity of strong clinical baseline. This suggests that all necessary information is not routinely extracted from WSIs, which advocates for the existence of unknown biomarkers. Further, it implies that diagnosis slides themselves contain enough information to predict NACT response. Last, but not least, this algorithmic approach seems to solve this problem with better performance than the traditional clinicopathological criteria.

Collaboratively trained WSI-based DL models generalize much better than their local counterparts, not only in internal cross-validation but also on a test set held out from the train set. We stress that this result still holds even when tested in very unfavorable conditions, in a setting where the total number of samples is of the same order of magnitude as the local number of samples [54]. It is also important to note that even on the sanctuarized test set, the federated WSI-based model maintains its superior performance overall. This shows the complementarity of the signals found in both cohorts, as evidenced by Fig. 2, and hints at a potential prospective usage.

The main limitation of our study, and simultaneously its strength, stems from grouping all TNBC together independently of their molecular subtypes. It is known that TNBC comprises at least 6 molecular subtypes [55] and that treatment effects should be investigated from within sub-groups [56]. Our model indeed underlines that such subgroups (Basal-like and Immunomodulatory versus LAR) were important in predicting the response and it is possible that finer grained analysis would be even more performant. Indeed it has been shown before that models can extract molecular information from WSI [57]. However, this information is not always available in cancer centers nor is there a definitive agreement on sub-types classifications for TNBC, which limits the deployment of specialized models. This makes our models more general, but possibly hinders their performances. Furthermore, populations of a specific molecular subtype would necessarily contain fewer slides and even more unbalanced distributed datasets. Thus it is unclear whether we would see actual performance gains in data-intensive DL methods.

Further investigations could be done using DL to discover sub-groups of more homogeneous TNBC, so that patients could be automatically tagged and assigned a specialized model only using raw WSIs. However clustering methods in private distributed settings [58] are still in their infancy, and most algorithms require to communicate sensitive information such as features’ means, which makes them less amenable to the Federated Learning setting.

Another limitation of our work is that it only investigates either clinical data models or WSI-based models. However, multimodal data integration is known to have brought improvements in several ML applications [59]. We defer this multimodal investigation to future works.

Interpretability results show that models have re-discovered known biomarkers such as TILs without any tile-level annotations, and hints at potential new potential biomarkers, such as fibrosis. This interpretability also highlights the importance of stromal regions in prediction of response to NACT with a lot of significant criteria located in such regions : TILs, fibrosis, hemorrhagic suffusion. However, while our model does use those patterns to inform its prediction, a more quantitative study to assess the direct impact of each criteria would be needed to validate those biological insights. Future works could go beyond this proof of concept, for instance, by measuring the univariate AUCs of the presence of each criteria with the response to NACT. Nevertheless, this analysis would be difficult to carry out, as annotating the newly found criteria at the tile level for 500 WSIs is extremely time-consuming for pathologists and is prone to inter-observer variability.

In conclusion, although it is apparent that the scientific community does not yet fully understand all factors involved in the response to NACT for early TNBC, we show the potential of data-driven methods for making progress. WSI-based models could be effective replacements for clinical models heuristics for the prediction of the response to NACT in the absence of better alternatives. Moreover we demonstrate in this pioneer multi-centric proof of concept that *real-world* Federated Learning allows to train DL models that are overall more robust and transferable, as evidenced by their improved performance. Such models, easier to use and more accurate, also provide interpretability heatmaps highlighting known biomarkers such as TILs and apocrine tumor cells [56], and generating hypotheses on potential new biomarkers such as discohesive tumor cells and fibrosis. Therefore, we advocate for an extensive testing of the performance of our Federated WSI-based models on other cohorts so that they can eventually be adopted in modern clinical practice as a support to existing heuristics until the next research breakthrough. More generally, this study showcases that Collaborative Federated Learning of interpretable Deep Learning models is a promising avenue in order to make novel discoveries in complex diseases such as Breast Cancer, as the heterogeneity of the related cohorts always offer a more accurate and complete picture than siloed studies.

## 5 Methods

### 5.1 Cohorts construction

#### Patient pathway

In both comprehensive cancer centers under study, Hospitals A (Centre Léon Bérard) and B (Institut Curie), giving rise to Cohorts A and B, respectively, we consider the patient pathway illustrated in Fig. 1 (a), that we now detail. Early TNBC gets diagnosed and relevant clinical information is extracted, including side of the diseased breast, age, body mass index and menopause status. Subsequent body CT scan and breast MRI provide the clinical TNM status of the cancer (cTNM) [60, 61]. The diagnostic biopsy is realized to extract the remaining relevant clinical variables mentioned in Fig. 1 (a). TILs is annotated in both centers using the same standard methodology described in [62]. It can happen that the HER2 status is equivocal in Immunohistochemistry: when this occurs, the Fluorescence In Situ Hybridization (FISH) technique [63] is applied to disambiguate. After NACT, at surgery the breast and the axillary lymph nodes are probed to measure the treatment effectiveness based on the Residual Cancer Burden (RCB).

#### NACT response

Responders are split into two groups according to a binarization of their RCB status after treatment. Details about the binarization of RCB to obtain the histological NACT response are provided in Tab. 4 in the Supp. Mat.

#### Dataset

After selecting records matching this pathway in each hospital, we end up with 102 slides in Cohort A associated with 99 patients and 427 slides in Cohort B associated with 420 patients. We treat each slide as an individual data point, but stratifications are done respecting the patient information. The complete list of clinical variables available in cohorts A and B are provided in Tab. 4 in the Supp. Mat.; the sets of variables available in the different medical institutions differ but their intersection is non empty. Both institutions are located in the same country although distant geographically.

#### WSI details

Slides in both cohorts were prepared onsite at their respective institutions with similar but slightly different staining protocols. Cohort A uses Hematoxylin Phloxine Saffron (HPS) and Cohort B uses Hematoxylin Eosin Saffron staining (HES) [64]. These stainings differ from traditional HE staining [64] by coloring the connective tissues differently than muscles and cytoplasm. Institutions also differ by the fixative and the scanners they use to produce WSI: Bouin’s fixative or formol [65] are systematically used in Cohort A and AFA(Alcohol Formalin Acetic acid) and/or formol [65] in Cohort B. Regarding scanners, in Cohort A, a Leica APERIO AT2 scanner was used producing SVS images, in Cohort B two scanners were used at different periods of time: a Philips Ultra Fast Scanner 1.6 RA and a Hamamatsu Nanozoomer S360 producing TIFF and NDPI images. In both centers, slides were scanned with up to x40 magnification but in practice we only used x20 zoom-level in our experiments (see Section 5.3) as we find it provides the best trade-off between the size of the context, the level of details of the tiles and the computational burden of the method, corroborated by the literature [66, 30].

#### Train-test dataset split

We reserve approximately 20% of the slides in each center for a sanctuarized test set where we evaluate only once the final models, and use the remaining ones for cross-validation purposes. In Cohort A, we thus set aside 20 slides and, in Cohort B, 60 slides. The purpose of this split is to provide a better generalization study of the developed ML models.

#### Cross-validation split

In order to develop our ML models, we split the aforementioned training set into 4 cross-validation folds on each center, stratifying on each center by patient and response to treatment, repeated independently 5 times. The folds are aligned across the two centers, so as to end up with 20 multi-centric training and validation sets. All results of all models are presented on the 20 distributed folds except when explicitly stated.

We also explored different intra-center subgroups based on the retrospective RCB status or the grade keeping only extremal values. However no significant performance effect was observed probably due to the small size of the resulting subgroups. Therefore, we kept the focus of our analysis on the original groups.

### 5.2 Analysis of diagnosis variables and NACT response

#### Univariate analysis

In Fig.2 (a), the AUC of each variable used to predict the NACT response is computed on the train set with 5 repetitions of 4-fold cross-validation, as well as the AUC of the opposite of the variable and the NACT response. We select the variable or its opposite depending on whether it has an average AUC to predict NACT response larger or smaller than 0.5, in order to report only AUCs larger than 0.5. We thus end up with 20 AUC values whose mean is larger than 0.5. The mean of these values is denoted in the bar height in Fig.2 (a). For each variable, the 95% confidence interval is computed with 1000 individual bootstraps of 20 values with replacement of these 20 AUCs.

#### Multivariate analysis

Logistic regression models’ training is described in Sec. 5.4. The AUC reported in Fig 2 (b) is computed on the validation sets. On the same figure, the box plots are computed as follows: the central bar denotes the mean, the edges of the box the first quartile and the last quartile, and the inter-whiskers distance is equal to 1.5 times the interquartile range, centered around the median.

### 5.3 WSI-based Models

#### 5.3.1 Slides Preprocessing

We first segment the matter on the WSI using a matter-segmentation U-Net [67] trained on a private dataset. We then tile the extracted matter using 224×224 tiles at zoom-level 20x. This results in a variable number of tiles for each slide depending on the amount of matter found on the slide. We finally rely on a Imagenet-pretrained ResNet-50 [68], truncated until its penultimate layer, to extract a feature of size 2048 for each tile. Each slide is eventually represented by a bag of such features.

The aforementioned pipeline above has proved sufficient to reach good performance, but could be improved in several directions listed below.

##### Normalization

Slide normalization in Federated-Learning is a relatively uncharted topic [30] because image-level normalization techniques [69, 70, 71] are difficult to apply as they require centers to agree on a common template. In this work, we make the first order approximation that Imagenet features alone allows to reduce the variability between the centers enough to successfully train a DL model across multiple institutions as evidenced in [72].

Recently, model-level normalization in Federated Learning has emerged [73] thanks to the Batch-Normalization layers [74] used in some of the Deep Learning models. However, as our models already transfer well without Batch-Normalization we devote this approach to future works.

##### Self-Supervised Learning on Histology Data

We also explored the extraction of features from a deep network pretrained on histology slides from TCGA-COAD [75] with a state-of-the-art self-supervised method [76] but did not observe any performance gain locally.

#### 5.3.2 Weakly Supervised Model Training

We use the Multiple Instance Learning (MIL) paradigm [77] to train models on labeled bags of features. We experiment with different aggregation functions and models: min-max based aggregation using Chowder [66] and Weldon [78], learnable pooling with DeepMIL [79] and logistic regression on the mean features called MeanPool [66], which all are standard approaches for weakly-supervised WSI classification.

To find appropriate hyperparameters for each model, we perform one 4-fold cross-validation grid search locally on Cohort B, which is the most difficult Cohort. Then, we evaluate the performances of these models on the selected hyperparameters on five times 4-fold cross-validations on each Cohort. Results are presented in Table 1 in the Supp. Mat. We observe that the MeanPool models perform better overall. Thus, we select this architecture for our experiments. The studied hyperparameters for the different models include the batch size, L1, L2, regularizations on the weights and the use of a locally trained autoencoder. The final MeanPool model does not use any autoencoder nor regularization and is trained with a batch-size of 16 by the ADAM optimizer [80] with a learning rate of 0.001.

#### 5.3.3 Collaborative Distributed Training

##### Ensemble

The Ensemble method consists in independently training local models and aggregating the predictions of the models proportionally to the number of samples in each cohort. More precisely, for a given WSI sample *x*, if *f*_*A*_ denotes the model trained locally on Cohort A and *f*_*B*_ is the model trained locally on Cohort B, as described in Sec. 5.3.2, then the prediction *y*_pred_ for the Ensemble strategy is given as *y*_pred_ = 0.18 · *f*_*A*_(*x*) + 0.82 · *f*_*B*_(*x*). The rationale behind these aggregation weights is both to provide a final predictor which is balanced with respect to the domain of origin of the samples, and to ensure a fair comparison with federated learning approaches, which use the same mixing weights for updates.

##### FL epoch selection

The FL strategies all use the same number of local minibatch updates in both centers. Minibatch size is set to 16 in all cases. Whatever the frequency of communication rounds, the total number of such local batch updates is defined with respect to the largest cohort size (Cohort B), so as to achieve 30 epochs on this cohort.

##### Federated Averaging (FedAvg)

FedAvg [42] consists in training locally the WSI-based models and periodically averaging their weights throughout training. Both cohorts take part in each synchronized round. We use *E* = 50 local batch updates before averaging in order to reduce communication costs, and compute the number of rounds as mentioned in the previous paragraph. As in the local case, we use the ADAM optimizer with a learning rate of 0.001. Weight averaging is done proportionally to the number of samples in each cohort, with the same mixing weights as for the Ensemble strategy prediction, which is standard for Federated Averaging [42].

##### Scaffold

For Scaffold [53] trainings, we follow the same exact setting as for FedAvg, i.e. we use *E* = 50 local batch updates and the same number of rounds, learning rate and optimizer. Regarding Scaffold-specific hyperparameters, we use a server learning rate of 1.0, and the update rule II, as defined in the original article. Note that in contrast to this work, [53] use traditional Stochastic Gradient Descent and not ADAM.

### 5.4 Clinical Data Models

#### Data preprocessing

To handle missing data, we use the most frequent levels found in the training set. Categorical features are dummified using a drop-last policy, i.e. for each variable with *k* levels we create *k-* 1 one-hot variables. For Logistic regression with ElasticNet, we scale the variables in each center, and use the same scaling parameter in both centers so as to be able to compare the magnitudes in Fig. 2 (b).

#### Model cross-validation

For each model type, we cross-validate the best hyperparameters. Cross-validation is performed independently on each Cohort As we don’t look at inter-center performances for the clinical models. The grid we use in our experiments can be found in Table 2 in the Supp. Mat.

#### Models used and associated hyperparameters

We study 3 types of clinical data models: CART [81], XGBoost [82] and Logistic regression with ElasticNet [51]. For CART, the only hyperparameter is the maximal depth of the tree. For XGBoost, the hyperparameters are the maximal depth of the constructed tree and step-size shrinkage *η*. For ElasticNet, the hyperparameter is the strength *C* of the ElasticNet penalty, with an *α* ratio between L1 and L2 of 1.

### 5.5 Interpretability

For the interpretability results, we extract the 50 highest and lowest scores in each cohort’s test set with a maximum of 5 tiles per slide using the scores given by the best Federated Learning model. In order to obtain a tile-level score, we just apply the model to each tile and remove the averaging step. We obtain 200 tiles that we shuffle and give to two pathologists to annotate according to a predefined list of criteria in a blinded fashion with respect to the tile score. The precise list of criteria is given in Supp. Mat. F.

### 5.6 Statistics and reproducibility

For all *p*-values we fix the significance threshold to 0.05.

#### Diagnosis variables and NACT response

For Fig. 2 (a) we use empirical *p*-values by bootstrapping the cross-validation score of a random model 10,000 times on both cohorts against the cross-validation score of each variable in a one-sided test. *p*-values are then corrected using the Holm correction [83] to limit false discovery rate. We emphasize that models are not trained per se, as we use the raw variables, but we still use folds to facilitate comparisons with ML models and to select the direction of each variable to use as predictor. Similarly, both Logistic Regression models from Fig. 2 (b) are evaluated against random model performances.

#### WSI-based models to predict NACT response

For Fig. 3 (a) to compute AUC comparison *p*-values across the same folds we use the one-sided Wilcoxon signed rank test [84]. In the case of Fig. 3 (b) since we only have point estimates we bootstrap 10,000 times groundtruth-prediction pairs from all models and compute a one-sided Z-test on the AUC difference.

#### Interpretability of WSI-based model

For Fig. 4 (b), we use the two-sided exact Fisher Test [85] assuming a Bernoulli distribution of the presence/absence of a given criteria to compute *p*-values comparing tile count in both min and max tiles.

## Data Availability

Data is available upon reasonable and GDPR compliant request from Institut Curie and Centre Léon Bérard

## 6 Acknowledgements

This project is supported by Bpifrance as part of the “Healthchain” project, which resulted from the “Digital Investments Program for the major challenges of the future” RFP. As part of the “Healthchain” project, a consortium coordinated by Owkin (a private company) has been established, including the Substra association, Apricity (a private company), the Assistance Publique des Hôpitaux de Paris, the University Hospital Center of Nantes, the Léon Bérard Center, the French National Center for Scientific Research, the École Polytechnique, the Institut Curie and the University of Paris Descartes

## 7 Author Contributions

P.-E.H. and G.B. conceived the idea of the paper with the help of Al.L. and reviewed the medical content of the paper. J.O.d.T., C.B., M.A, Ch.M., B.S., E.W.T. implemented the analysis and wrote the article, while G.W. and M.Z. supervised the methodology. C.J., Ar.L. and M.M. were in charge of data curation and harmonization in both centers. J.Ge. and G.B. also annotated the pathology images and provided the interpretations of the histological patterns found. C.G., K.M., I.D., Ca.M, T.D. and J.Gu. were in charge of the federated network between the hospitals (IT infrastructure and software backend using substra). M.G, A.-L.M., E.B., T.D. and Al.L. managed the collaboration.

## 8 Code and Data availability

Data underpinning this study are under restricted access and are not publicly available. Data might however be made available upon reasonable and GDPR compliant request from Centre Léon Bérard and Institut Curie. All federated learning experiments relied on Substra, an open source software available at https://github.com/SubstraFoundation/substra. The scripts to run these experiments are not public, but detailed information in the methods allows to reproduce them.

## 9 Ethical Approval

The IRB of Institut Curie (Comité de Revue Institutionnelle - CRI Data) and the IRB of Centre Léon Bérard (Artificial Intelligence Project Evaluation Board) both validated the ethics of the current study inside the HealthChain project.

