## Supplementary Material for "Collaborative Federated Learning behind Hospitals’ Firewalls for Predicting Histological Response to Neoadjuvant Chemotherapy in Triple-Negative Breast Cancer"

#### A ML models fine-tuning and hyperparameters

| | Cohort B (1 times 4-fold) | Cohort B (5 times 4-fold) | Cohort A (5 times 4-fold) |
|---|---|---|---|
| MeanPool | $\mathbf{0.6091 \pm 0.0486}$ | $\mathbf{0.5661 \pm 0.0544}$ | $\mathbf{0.7135 \pm 0.0779}$ |
| Weldon | $0.5587 \pm 0.0600$ | $0.5553 \pm 0.0520$ | $0.6681 \pm 0.1047$ |
| Chowder | $0.5881 \pm 0.0240$ | $0.5479 \pm 0.0500$ | $0.5896 \pm 0.1438$ |
| DeepMIL | $0.5827 \pm 0.0300$ | $0.5420 \pm 0.0612$ | $0.6775 \pm 0.0656$ |

**Table 1.** Mean AUCs reached by selected hyperparameters from one 4-fold cross validation grid search on Cohort B.

| Model | Hyperparameters Grid |
|---|---|
| CART | $D = [1, 2, 3, 4, 5, 6, 7, 8]$ |
| XGBoost | $D = [1, 2, 3, 4, 5, 6, 7, 8]$, $\eta = [0.001, 0.005, 0.01, 0.05, 0.1, 0.5]$ |
| ElasticNet | $C = [1., 10., 100., 1000., 0.1, 0.01, 0.001]$ |

**Table 2.** Hyper-parameters grid

| Indicator | | Cohort A | Cohort B |
|---|---|---|---|
| **# slides** | **Training** | 82 | 367 |
| | **Test** | 20 | 60 |
| | **Total** | 102 | 427 |
| **Pathology Data** | | Bouin Fixative until 2010 then Formol<br>TILs annotated by one pathologist<br>a posteriori | AFA Fixative before 2016 then Formol<br>TILs annotated by multiple pathologists<br>through the years, reflecting clinical practice |
| **Specific clinical data** | | Sum of Tumor Sizes, Principal Tumor Sizes, pM | KI67, FISH, insitu carcinoma, mitosis, necrosis |
| **Clinical data status** | | No missing data | Some missing data (9.22% on average on common variables) |

Table 3: Overview of the cohorts

### B Cohorts and variables description

| Clinical Feature | Description | Unit | Availability |
|---|---|---|---|
| *Patient Information* | | | |
| **Sex** | Sex of the patient | M<br>F | A/B |
| **Weight** | Patient weight at time of diagnosis. | kg | A/B |
| **Age** | Patient age according to one of four pre-specified age ranges. | (0-25]<br>(25- 50]<br>(50- 75]<br>(75- 100] | A/B |
| **Body-Mass Index (BMI)** | Patient BMI at the time of diagnosis. | kg / m2 | A/B |
| **Menopause Status** | Patient Menopause status at the time of diagnosis. | pre<br>post<br>unspecified | A/B |
| *Pre-Treatment Measurements* | | | |
| **Biopsy Year** | The year at which the diagnosing biopsy was collected. | year | A/B |
| **Side** | On which breast was the diagnosis made. | left<br>right | A/B |
| **Insitu Carcinoma** | Whether the biopsy contains insitu carcinoma. | Yes<br>No | B |
| **Multifocal Tumor** | The cancer is multifocal when there are at least two separated lesions. | Yes<br>No | A/B |
| **cT** | PRE-treatment clinical TNM status.<br>Categorization of primary tumor. | Tx/Na<br>T0<br>Tis<br>T1<br>T2<br>T3<br>T4a<br>T4b<br>T4c<br>T4d | A/B |
| **cN** | PRE-treatment clinical TNM status.<br>Categorization of spread to regional lymph nodes. | Nx/No<br>N1<br>N2<br>N3 | A/B |
| **cN** | PRE-treatment clinical TNM status.<br>Presence of distant metastasis. | M0<br>M1 | A/B |
| *Pre-Treatment Pathology Information (pathologist assessment)* | | | |

| | | | |
|---|---|---|---|
| **TILs** | Percentage of tumor-infiltrating T-lympocytes observed in the pre-treatment histology; given as a continuous value. Assessed by one or multiple pathologists, depending on the cohort. | [0-100] percentage | A/B |
| **EE grade** | Elston and Ellis Histological grade. | 1<br>2<br>3 | A/B |
| **Histo. Subtype** | Classification of which of the three primary breast cancer subtypes the diagnosis belongs to. | Invasive Carcinoma<br>ISC- Ductal<br>ISC-Lobular | A/B |
| **PR** | The progesterone-receptor-positive (PR+) status of the tumor, graded using IHC. | Uninterpretable<br>Negative or (0, 10%]<br>Negative or (10, 50%]<br>Negative or (50, 80%]<br>Negative or (80, 100%] | A/B |
| **ER** | The estrogen-receptor-positive (PR+) status of the tumor, graded using IHC. | Uninterpretable<br>Negative or (0, 10%]<br>Negative or (10, 50%]<br>Negative or (50, 80%]<br>Negative or (80, 100%] | A/B<br>A/B |
| **HER2-IHC** | Result of Immunohistochemistry-based test for HER2 in pathology. A value of +++ represents a confident HER2 positive measurement, while ++ is borderline, and 0/+ represents an HER2-negative test. | 0/+<br>++<br>+++ | A/B |
| **FISH** | Fluorescence In Situ Hybridization (FISH), a more accurate measurement which can be employed to corroborate the IHC-based HER2 test for the HER2 status of tumors cells. | Amplification +<br>Absence of Amplification - | B |
| **KI67** | Ki-67 Labelling Index. Measurement of the presence of Antigen Ki-67 (Marker of Proliferation, Ki-67), a nuclear protein associated with cellular proliferation. | [0-100] percentage | B |
| **Presence of Vascular Emboli** | | Present<br>Not Present | A/B |
| **Necrose** | Whether or not necrotic tissue was observed in pathology. | Observed<br>Not Observed | |
| **Mitosis** | | 1<br>2<br>3 | B |
| *Post-Treatment Measurements* | | | |
| **pT** | POST-treatment pathological TNM status. Categorization of primary tumor | Tx/Na<br>T0<br>Tis<br>T1<br>T2<br>T3 | A/B |

| | | | |
|---|---|---|---|
| | | T4a<br>T4b<br>T4c<br>T4d | |
| **pN** | POST-treatment pathological TNM status. Categorization of spread to regional lymph nodes. | Tx/Na<br>Nx/No<br>N1<br>N2<br>N3 | A/B |
| **pM** | POST-treatment pathological TNM status. Presence of distant metastasis. | M0<br>M1 | A |
| **RCB** | Combination of measurements of tumor, tumor bed, and metastasis present in axillary lymph nodes | continuous index | B |
| **RCB Class** | Severity scoring of the continuous RCB measurement. Scoring is calculated according to predetermined thresholds (0, 1.36, 3.28). | 0<br>I<br>II<br>III | B |
| **Principal Tumor Size** | Direct measurement of the principal tumor after treatment. | mm | A |
| **Sum of Tumor Sizes** | Sum of sizes of all identified tumors after treatment. | mm | A |
| **NACT Histological Response** | **Study Endpoint.** Classification of whether or not the patient responded to neo-adjuvant chemotherapy: thresholding of RCB Class. | RCB Class 0 – Complete<br>RCB Class I II III – Incomplete | A/B |

Table 4: Description and categorization of available per-patient clinical data across all cohorts. Cells in yellow indicate admission measurements which serve as criteria for admission into the study cohorts.

#### C Analysis of differences in Cohorts A and B

We compare the distribution of each common clinical variable between both cohorts in order to track the origin of the differences in the predictions' performances. We display Fig. 5 the univariate histograms of the most discriminative variable between the two cohorts: age, EE grade and cT status. By inspecting the oldest group of patients in Cohort B, we see that among 14 patients only 1 had a positive complete response to NACT, in the rest of the cohort there are 51.2% good responders. This result is expected because older patients are more at risks.

These comparisons of clinical data distributions between the two cohorts demonstrate that despite the alignment of data between the two cohorts, the cohorts are still heterogeneous. In addition, we note that in Cohort B, biopsies before 2017 are from a retrospective cohort, whereas biopsies received after 2017 were not curated. However, the original source of disparities between both populations is still relatively unknown as it does not seem to appear strongly in the distributions of the extracted clinical variables.

#### D TNBC studies patient count

#### E Clinical Practice: Model's Confusion Analysis

For each trained model, we complement Fig. 3 b) by providing confusion matrices, for the best model of each type in each cohort, by optimizing the decision threshold in order to maximise the specificity while keeping a maximum of 25% false negative patients.

#### F Annotation Pathology Criteria

In Table 5, each criterion is associated with the values it can take whether it is percentage, discrete levels (grading or presence/absence of specific levels), or boolean (presence/absence). Scoring and annotations of the architecture, atypia and mitosis in the first rows of the table refer to the 3 components of the EE tumor

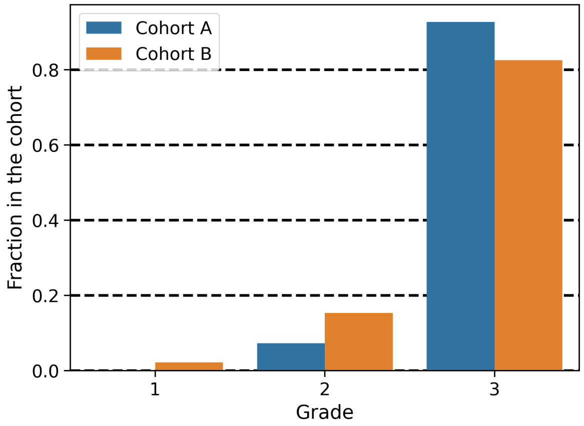


**(a)** EE grade distribution

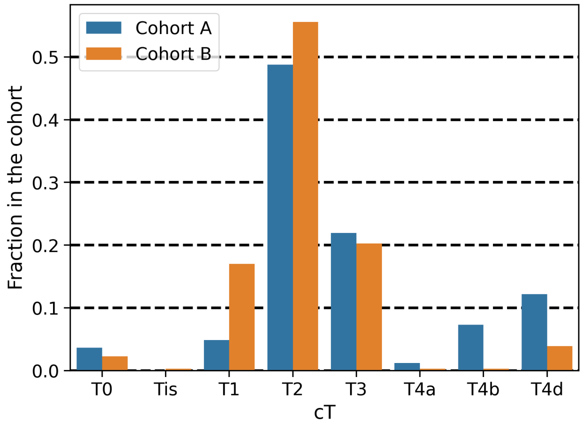


**(b)** cT distribution

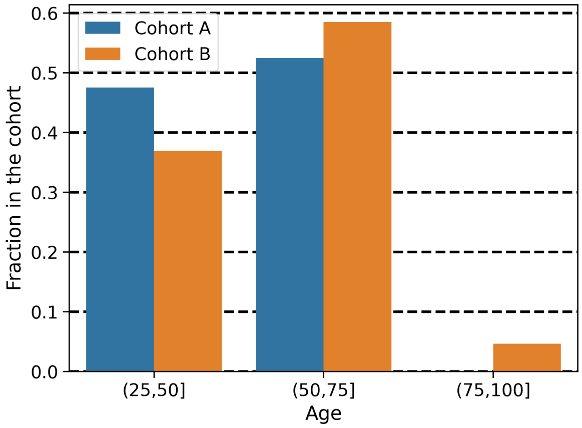


**(c)** Age distribution

**Fig. 5.** Distribution of the most discriminative variables between the tow cohorts: EE grade, cT and age in each cohort

grade. Regarding the Dominant architecture annotations, we call fibrosis the results from the deposition of a cross-linked collagen matrix by cancer-associated fibroblasts (CAFs). We also call discohesive tumor cells, the classic histological micro pattern of ILC (Invasive Lobular Carcinoma) defined as "characterized by proliferation of non-cohesive small cells individually dispersed in fibrous connective tissue or organized in single-file linear cords invading the stroma" [88].

**Fig. 6.** TNBC studies patient counts.

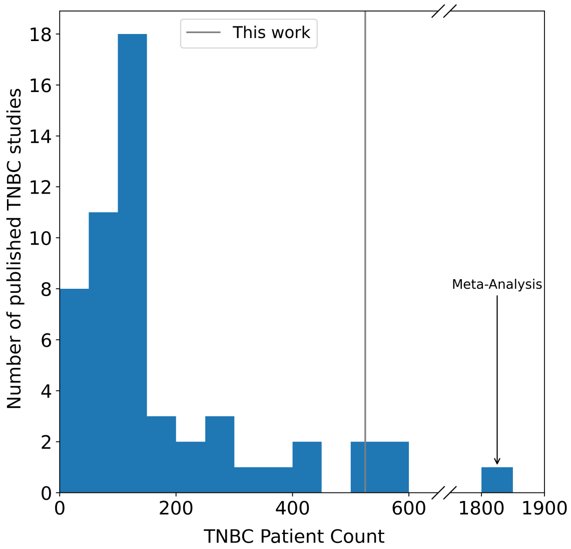


Patient count in recent TNBC studies following a comprehensive bibliographic search. We extracted 54 articles from either the survey from Bianchini et al. [86] or from our article using all citations that contain the keywords ”triple negative”, ”triple-negative” or ”TNBC” and from which the number of patients could be extracted upon reading. Out of 55 TNBC studies including ours, our article is in the top 10% in terms of patient count. Median patient count is 119, with 25-75% quantiles 82-207. The study on the far right from Couch et al. [87] with 1,800 patients is a meta-analysis of multiple other studies.

| | | |
|---|---|---|
| **Epithelioid Structures** | Tumor cellularity | presence/absence |
| | Architecture | 1 to 3 |
| | Dominant Architecture | solid, span, discohesive cells, cribriform, tubes, micropapillary |
| | Atypias | 1 to 3 |
| | Mitosis | presence/absence |
| | Dominant Aspect of the Tumor cells | pleomorphic, fusiform, monomorphic, cytoplasm |
| | visible nucleoli | Yes, No |
| | Abundance of Cytoplasm | Yes, No |
| | Mucus | presence/absence |
| | Necrosis | presence/absence |
| | Tumorous Emboli | presence/absence |
| | Epidural sheathing | presence/absence |
| **Stromal Structures** | Stroma | presence/absence |
| | TILs | % of the tile area |
| | Other inflammatory cells | neutrophils, macrophages, nothing |
| | Fibrosis | presence/absence |
| | Elastose | presence/absence |
| | Calcifications | presence/absence |
| **Normal Structures** | Normal Glands | presence/absence |
| | Vascular Structures | presence/absence |
| | Hemosidirin | presence/absence |
| | Hemorrhagic Suffusion | presence/absence |
| | Mantle tissue (adipocytes, fibrosis) | |
| **Artifacts** | detachment, out of focus, wrinkles, No artefact | Yes, No |

Table 5: Tiles Pathology Criteria

**Fig. 7.** Confusion Analysis of models.

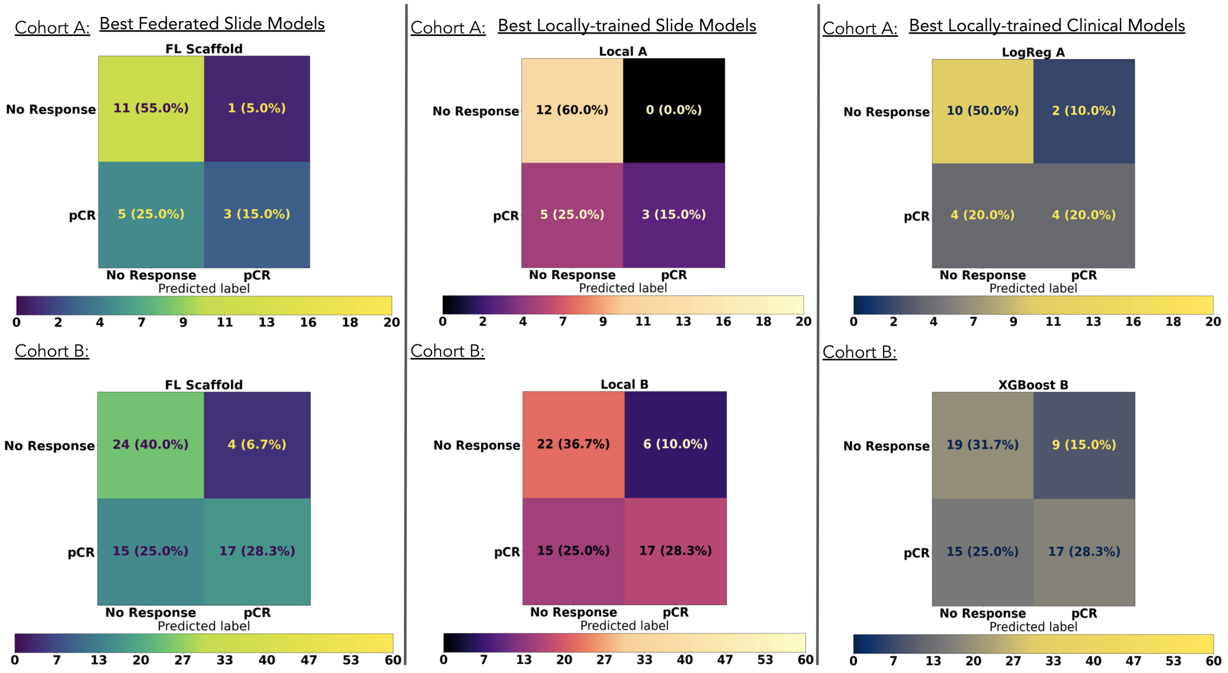


We display on the sanctuarized test sets, the confusion matrices obtained by choosing the classification threshold maximizing specificity while producing in each Cohort A maximum of 25% false negatives patients. For clinical practice false negatives is the worst outcome possible: patients that could be cured by the standard treatment are oriented towards second line treatments potentially more toxic and hazardous. For each model category we choose the best model according to its AUC point estimate.
